# Integration of a Molecular Prognostic Classifier into the Ninth Edition TNM Staging of Lung Adenocarcinoma

**DOI:** 10.64898/2026.02.17.26346484

**Authors:** Hanie Abolfathi, Fabien C. Lamaze, Sébastien Renaut, Michaël Maranda-Robitaille, Kelly-Ann Pellerin, David Joubert, Victoria Saavedra Armero, Nathalie Gaudreault, Dominique K. Boudreau, Michèle Orain, Patrice Desmeules, Andréanne Gagné, Yasushi Yatabe, Yohan Bossé, Philippe Joubert

## Abstract

**Introduction:** Despite advancements in non-small cell lung cancer (NSCLC) management through the use of molecular biomarkers, the recently introduced 9^th^ edition of the TNM staging system remains based exclusively on anatomic descriptors, with no consistently demonstrated improvement in risk stratification for early-stage disease. This study explores the integration of a molecular prognostic classifier into the conventional TNM staging system.

**Methods:** We analyzed 502 patients with stage I–III lung adenocarcinoma (LUAD) who underwent surgical resection with tumor-based gene expression profiling at the Quebec Heart and Lung Institute. A molecular prognostic classifier was developed and integrated into the 9^th^ edition TNM staging system to generate a novel model (TNMEx). Prognostic performance was compared with the 8^th^ and 9^th^ TNM editions using prognostic discrimination and reclassification metrics. External validation of the molecular classifier was performed in 271 LUAD cases from The Cancer Genome Atlas (TCGA). An independent cohort of 606 resected LUAD patients from the National Cancer Center Hospital (Tokyo) was used to externally compare the prognostic performance of the 8^th^ and 9^th^ TNM staging systems in the absence of molecular data.

**Results:** The molecular prognostic classifier was developed based on the expression levels of 26 prognosis-associated genes, weighted by their corresponding coefficients. The classifier was subsequently integrated into the 9^th^ edition TNM staging to generate the TNMEx model. The TNMEx system demonstrated superior prognostic performance, achieving a higher concordance index (C-index = 0.72) compared to the 9^th^ edition TNM (C-index = 0.65, p=0.006). Moreover, TNMEx significantly improved patient risk reclassification compared to both the 8^th^ (net reclassification improvement [NRI] = 0.27, integrated discrimination improvement [IDI] = 0.04) and 9^th^ editions (NRI = 0.40, IDI = 0.05), underscoring its superior ability to stratify outcomes. The 8^th^ and 9^th^ editions showed only limited improvement in overall prognostic accuracy and risk stratification, as reflected by their relatively modest C-index values (0.62 and 0.65, respectively) and minimal reclassification gains (NRI = −0.06, IDI = 0.003).

**Conclusions:** Incorporating a molecular-based prognostic model significantly enhanced the ability to recognize patients at high risk and to predict their survival outcomes more accurately than traditional TNM staging systems.

## 1. INTRODUCTION

Lung adenocarcinoma (LUAD) is the most prevalent subtype of non-small cell lung cancer (NSCLC), accounting for approximately 40% of all lung cancer cases worldwide [1–3]. Despite advances in the management of LUAD, it remains the leading cause of cancer-related mortality [4, 5]. The prognosis for LUAD remains poor, with a dismal 5-year survival rate of 10-22% [6]. Even in early-stage disease, around 13-23% of patients eventually relapse despite curative-intent surgery [7, 8].

The concept of cancer progression was first outlined in 1907 by Halsted et al. [9], who described solid tumors as initially growing in size, then spreading to regional lymph nodes, and eventually metastasizing to distant organs. Building on this understanding, Pierre Denoix [10] introduced the TNM staging system in 1953 to standardize the classification of tumors based on their size (T), nodal involvement (N), and presence of metastasis (M). Today, the TNM system remains a cornerstone in the clinical management of lung cancer [11, 12]. Although our knowledge of the molecular mechanisms underlying tumor biology has greatly advanced, the core descriptors-TNM staging, continue to serve as the foundation for prognosis and treatment decisions more than 70 years later.

One major drawback of the TNM staging system is its limited capacity to distinguish truly high-risk individuals, those with hidden metastases or aggressive tumor characteristics not reflected in current TNM parameters, from low-risk patients who are likely cured through surgery alone. This shortcoming contributes to significant mortality in early-stage disease and results in delayed or missed postoperative therapies for patients who later develop metastases [13].

Over the past ten years, the TNM staging has undergone three revisions, with the most recent 9^th^ edition officially implemented in 2024 [14–16]. Notable updates in the 9^th^ edition include splitting the N2 category into single-station and multistation N2 disease, as well as redefining the M1c category to differentiate between single-organ and multi-organ system metastases [15, 17].

Despite the comprehensive dataset used to develop the 8^th^ edition of the TNM staging system, external validation studies have shown limited prognostic performance, with overlapping survival between some adjacent stage groups even among patients with early-stage lung cancer [18–21].

Over the past decade, numerous investigations have shown that molecular biomarkers are critical determinants of tumor behavior and can serve as predictors of patient survival and responsiveness to particular systemic treatments [22–26]. Recent researches have suggested that integrating molecular features into the 8^th^ edition may enhance its ability to predict survival outcomes [13, 27]. Improving risk stratification in early-stage NSCLC is challenging, largely due to the constraints of using only the anatomical criteria of the TNM system. Further improvements in staging may be difficult without incorporating prognostic biomarkers that capture intrinsic biological features of tumor cells.

These findings suggest that enhancing prognostic accuracy will require the incorporation of tumor molecular features, a potential improvement that remains unexplored in the context of the newly revised 9^th^ edition. The next major advancement in lung cancer prognosis should focus on integrating molecular profiles to the conventional TNM staging systems, and leveraging the growing availability of personalized treatment [28, 29].

In our study, we hypothesized that the integration of a molecular prognostic classifier into the conventional TNM staging system could enhance risk stratification and improve the prediction of OS in patients with LUAD. To evaluate this hypothesis, we aim to develop a novel staging system that integrates tumor-based gene expression profiles into the 9^th^ edition TNM staging and assess its prognostic performance.

## 2. METHODS

### 2.1. Patient Cohorts

A total of 502 patients with LUAD who had undergone complete (R0) surgical resection at the Institut universitaire de cardiologie et de pneumologie de Québec - Université Laval (IUCPQ UL), Canada, between 2013 to 2023 and part of the LORD cohort [30–32] were included. Clinical, pathological, and molecular data were obtained from institutional records and the IUCPQ-UL Biobank.

An additional external validation cohort consisting of 271 LUAD cases from The Cancer Genome Atlas (TCGA) was used to confirm the model’s reproducibility and generalizability across populations. In addition, an independent external cohort of 606 patients with surgically resected LUAD from National Cancer Center Hospital (NCCH), Tokyo, Japan, was included to externally validate the prognostic performance of the 8^th^ and 9^th^ edition TNM staging systems. **Table 1** shows the clinicopathological features of the three cohorts, including the development and validation cohorts.

**Table 1:** Clinical and Pathological Characteristics of Patients in The LORD Development Cohort and The TCGA Validation Cohort.

| Characteristics | LORD cohort (N=502) | TCGA cohort (N=271) |
| --- | --- | --- |
| **Sex** |  |  |
| • Female | 275 (54.8%) | 152 (56.0%) |
| • Male | 227 (45.2%) | 119 (44.0%) |
| **Age at resection (year)** |  |  |
| • Mean (SD) | 64.9 (8.3) | 64.2 (9.0) |
| • Range | 26-83 | 33-87 |
| **Smoking Status** |  |  |
| • Ever | 472 (94.0%) | 253 (93.3%) |
| • Never | 30 (6.0%) | 18 (6.7%) |
| **Pathologic N** |  |  |
| • N0 | 395 (78.7%) | 187 (69.0%) |
| • N1/2 | 107 (21.3%) | 84 (31.0%) |
| **Pathologic T** |  |  |
| • T1 | 249 (49.6%) | 97 (35.8%) |
| • T2 | 196 (39.0%) | 146 (53.9%) |
| • T3 | 36 (7.2%) | 20 (7.4%) |
| • T4 | 21 (4.2%) | 8 (2.9%) |
| **TNM Stage** |  |  |
| • 1A1 | 8 (1.6%) | 4 (1.6%) |
| • 1A2 | 116 (23.1%) | 60 (22.1%) |
| • 1A3 | 90 (17.9%) | 16 (5.9%) |
| • 1B | 123 (24.5%) | 87 (32.1%) |
| • 2A | 45 (9.0%) | 28 (10.4%) |
| • 2B | 66 (13.1%) | 38 (14.0%) |
| • 3A | 52 (10.4%) | 33 (12.1%) |
| • 3B | 2 (0.4%) | 5 (1.8%) |
| **Tumor size (cm)** |  |  |
| • Mean (SD) | 2.9 (1.7) | 1.3 (0.56) |
| • Median (Range) | 2.5 (1.0-9.2) | 1.1 (0.2-3.2) |
| **Tumor location** |  |  |
| • Upper Lobe | 310 (61.8%) | 132 (48.7%) |
| • Lower Lobe | 155 (30.8%) | 66 (24.4%) |
| • Other | 37 (7.4%) | 73 (26.9%) |

Note: Tumor size refers to the invasive component of the tumor.
For the development cohort (LORD), TNM stage, refers to the 9<sup>th</sup> edition of the TNM staging, while for the validation cohort (TCGA), it refers to the 6<sup>th</sup>/7<sup>th</sup> editions.
*Italics* are used to indicate “age” and “tumor size” as continuous variables.

The inclusion criteria for both the development and validation cohorts were as follows: (1) histologically confirmed LUAD, (2) complete surgical resection with negative margins (R0), and (3) adequate tissue available for both histological and molecular analyses. Exclusion criteria included: (1) stage IV disease, (2) specific histological variants of LUAD (fetal, colloidal, enteric, and mucinous subtypes), (3) receipt of neoadjuvant therapy, (4) multifocal or synchronous tumors, and (5) combined carcinoma histology.

### 2.2. Histopathological Evaluation

Hematoxylin and eosin (H&E) stained sections from each case were collected and independently evaluated by thoracic pathologists (PJ, YY, PD, AG). Tumors were classified according to the 2021 grading framework proposed by the IASLC for non-mucinous lung adenocarcinoma (NM-LUAD) [33, 34]. Histological assessment involved documenting architectural growth patterns, the presence of spread through air spaces (STAS), lymphovascular invasion (LVI), visceral pleural invasion (VPI), as well as tumor dimensions and pathological stage based on the AJCC Cancer Staging Manual [15, 35].

Tumor grading was performed based on the 2021 International Association for the Study of Lung Cancer (IASLC) classification, [33, 34]. LUADs were categorized by estimating the proportion of six major histologic patterns, acinar, papillary, solid, lepidic, micropapillary, and complex glandular patterns (cribriform and fused glands), which together accounted for 100% of the tumor architecture [33, 34]. Tumor grade was assigned according to the predominant growth pattern and the presence or absence of high-grade patterns, in accordance with IASLC recommendations, classifying tumors as grade 1 (lepidic-predominant), grade 2 (acinar or papillary-predominant without high-grade patterns), or grade 3 (any tumor containing ≥ 20% high-grade patterns, including solid, micropapillary, or complex glandular patterns; including cribriform and fused glands) [33, 34, 36, 37]. TNM staging was determined based on the criteria established by the American Joint Committee on Cancer (AJCC) [15, 35].

5-year OS was calculated from the date of surgery to the date of death, last follow-up, or censored at 5 years. Both the 8^th^ and 9^th^ editions of the TNM staging system were applied to evaluate and compare their prognostic performance. All study procedures were approved by the institutional review board of IUCPQ-UL (ethics approval number: 2020-3352, 21871).

### 2.3. Molecular Prognostic Classifier, and Novel TNM Staging Development

Tumor RNA was extracted and subjected to library preparation, high-throughput sequencing, and transcript quantification; full methodological details are provided in the **Method S1**. We developed a molecular prognostic classifier and integrated into the 9^th^ edition TNM staging system to improve risk stratification and survival prediction in LUAD.

To develop a molecular prognostic classifier, we identified the most prognostically informative genes in tumor samples. Then a molecular risk score for each patient was calculated (for details of model training and feature selection, see section 2.4). Patients were subsequently categorized into low-, intermediate-, and high-risk groups based on tertiles of the molecular risk score: the lowest 33^rd^ percentile defined the low-risk group, the highest 33^rd^ percentile defined the high-risk group, and those in between were assigned to the intermediate-risk group.

Then the molecular risk grouping was incorporated into the 9^th^ edition TNM staging system to develop a novel integrated staging model, referred to as TNMEx, where “Ex” represents gene expression profiles. The reclassification process was guided by molecular risk group assignments: high-risk patients were up-staged by one TNM level, low-risk patients were down-staged by one level, and intermediate-risk patients remained in their original stage.

### 2.4. Statistical Analyses

Full details of the statistical analyses, including sensitivity analyses, model optimization procedures, and gene selection strategies, are provided in **Methods S2**. A molecular prognostic classifier was developed using Least Absolute Shrinkage and Selection Operator (LASSO) penalized Cox proportional hazards regression implemented with OS as the primary endpoint **(Methods S3)**. A molecular risk score was calculated for each patient by linearly combining the expression levels of selected genes weighted by their corresponding regression coefficients derived from the LASSO-Cox model **(Methods S4)**.

Survival analyses were performed using Kaplan–Meier methods and multivariable Cox regression models, adjusting for relevant clinicopathological covariates including age, sex, smoking status, tumor grade, tumor size, STAS, LVI, VPI, and TNM stage. To evaluate the added prognostic value of the molecular classifier, we compared the performance of the 8^th^ edition TNM staging, the 9^th^ edition TNM staging, and the integrated TNMEx model. Model performance was assessed using hazard ratios (HRs), concordance index (C-index), area under the curve (AUC), likelihood-ratio chi-square statistics, Akaike Information Criterion (AIC), Bayesian Information Criterion (BIC), and Nagelkerke’s R². Prediction accuracy was further evaluated using mean absolute error (MAE) and root mean squared error (RMSE), comparing Cox model–derived 5-year mortality probabilities with Kaplan–Meier–estimated 5-year observed mortality. Reclassification performance was assessed using Net Reclassification Improvement (NRI) and Integrated Discrimination Improvement (IDI) to quantify the improvement in risk stratification provided by the TNMEx model.

The LORD cohort (n = 502) was randomly divided into a training cohort (n = 350) and an independent testing cohort (n = 152) using a 3:1 ratio with balanced clinicopathological characteristics **(Table S1)**. The training cohort was used to develop the molecular prognostic classifier, whereas the testing cohort was used to evaluate its prognostic performance.

The TCGA cohort was used exclusively to validate the molecular prognostic classifier, whereas the NCCH cohort was used to independently evaluate and compare the prognostic performance of the 8^th^ and 9^th^ edition TNM staging systems, without application of the TNMEx model. Cases from the TCGA, and NCCH cohorts were selected using the same inclusion and exclusion criteria applied to the LORD cohort, as described in the Patient Cohort section. All statistical analyses were conducted using R version 4.5.1, with two-sided p-values < 0.05 considered statistically significant.

## 3. RESULTS

### 3.1. Patients Characteristics

In the whole LORD cohort, the median follow-up was 87.7 months. Patients had a mean age of 64.9 years at diagnosis (range, 26-83), and 54.8% were female. Most patients were ever-smokers (94.0%), and the median tumor size was 2.9 cm (range, 1.0-9.2 cm) **(Table 1)**.

Based on the 9^th^ edition TNM staging, 337 patients (67.1%) had stage I disease, 111 (22.1%) stage II, and 54 (10.8%) stage III (**Figure S1**). Tumor grading showed that 40 tumors (8.0%) were grade 1, 123 (24.5%) were grade 2, and 334 (66.5%) were grade 3. LVI, VPI, and STAS were present in 49.0%, 30.9%, and 46.6% of cases, respectively. By the end of follow-up, 224 patients (44.6%) had died, and using 5-year OS censoring, 137 patients (27.3%) had died, and the remaining patients were either censored or alive at the 5-year mark.

### 3.2. Incorporating the Molecular Prognostic Classifier into the TNM Staging

The LASSO penalized Cox regression, applied separately to the training set, identified a panel of 26 genes associated with patient survival. This diverse gene signature encompasses protein-coding and non-coding genes, reflecting the multifactorial nature of LUAD progression and survival. These genes are involved in key biological pathways relevant to cancer progression, such as cell proliferation and survival, immune evasion, stress response and metabolism, epithelial-mesenchymal transition, and therapy resistance. The description, and the functional roles of genes are available in **Tables S2**, and **S3,** respectively.

Each patient received a molecular risk score and was stratified into low-, intermediate-, and high-risk groups. Based on this classification, 167 patients were categorized as high-risk, 167 as intermediate-risk, and 168 as low-risk.

The 26-gene prognostic classifier was then applied to the testing cohort using the same LASSO-derived coefficients obtained from the training set. As illustrated in **Figure 2 (A-C)**, molecular risk stratification revealed significant differences in 5-year OS across TNM stages I (p < 0.0001), II (p = 0.036), and III (p = 0.048), with high-risk patients exhibiting substantially worse outcomes than those in the intermediate- and low-risk groups. These molecular risk groups were then integrated into the 9^th^ edition TNM staging system to make the TNMEx. This reclassification preserved the TNM hierarchy while allowing up-staging for high-risk and down-staging for low-risk profiles. Overall, 66.1% of patients were reclassified under the TNMEx system, with redistribution across all TNM stages **(Figure 1)**. The final TNMEx stage distribution was 316 in stage I, 110 in stage II, and 76 in stage III **(Figure S1)**.

**Figure 1:**
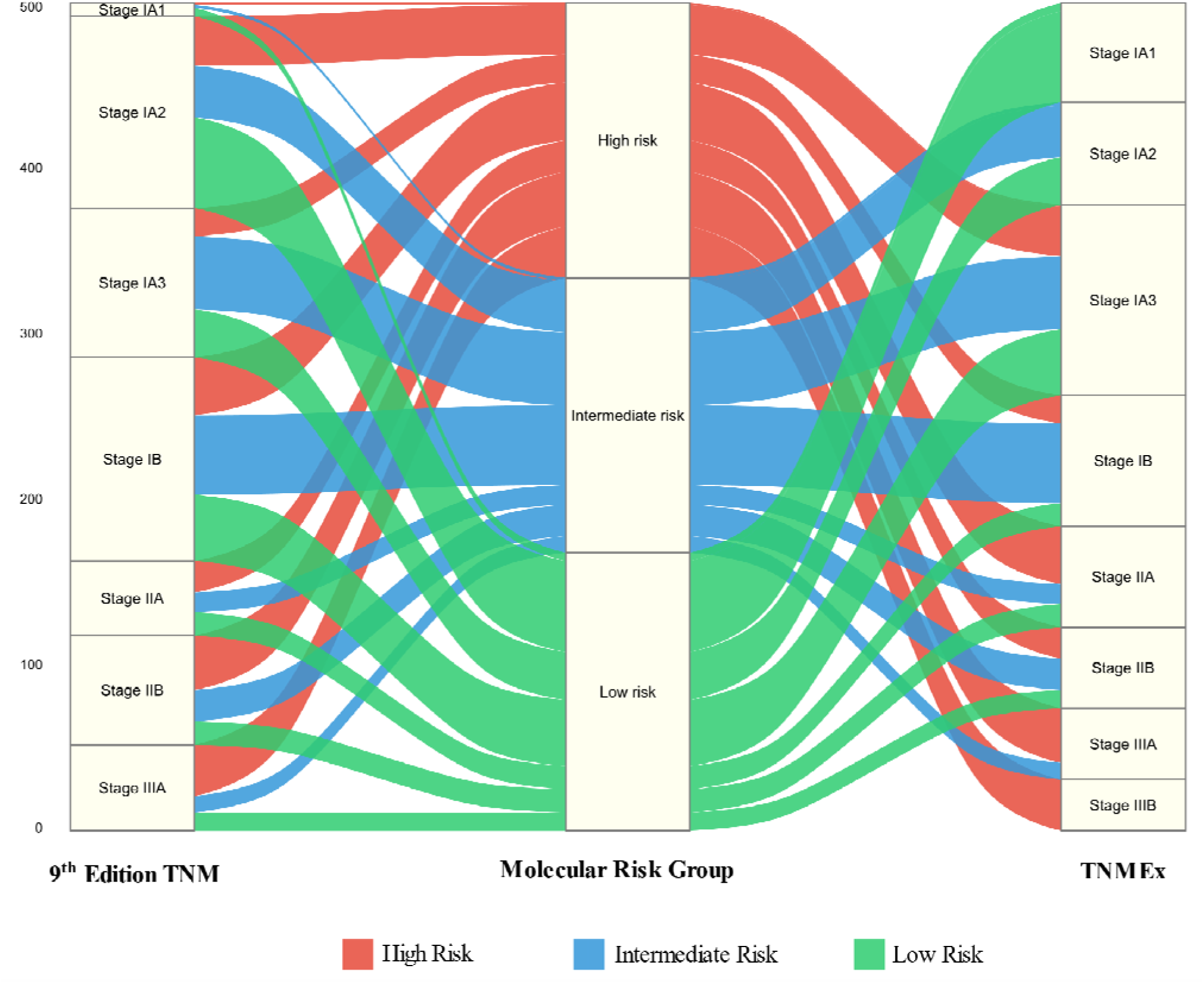
TNMEx Staging System in the Development Cohort (LORD, N=502). Note: In the TNMEx model, Ex refers to incorporated gene expression profiles to the overall structure of the 9^th^ edition TNM staging system.

**Figure 2:**
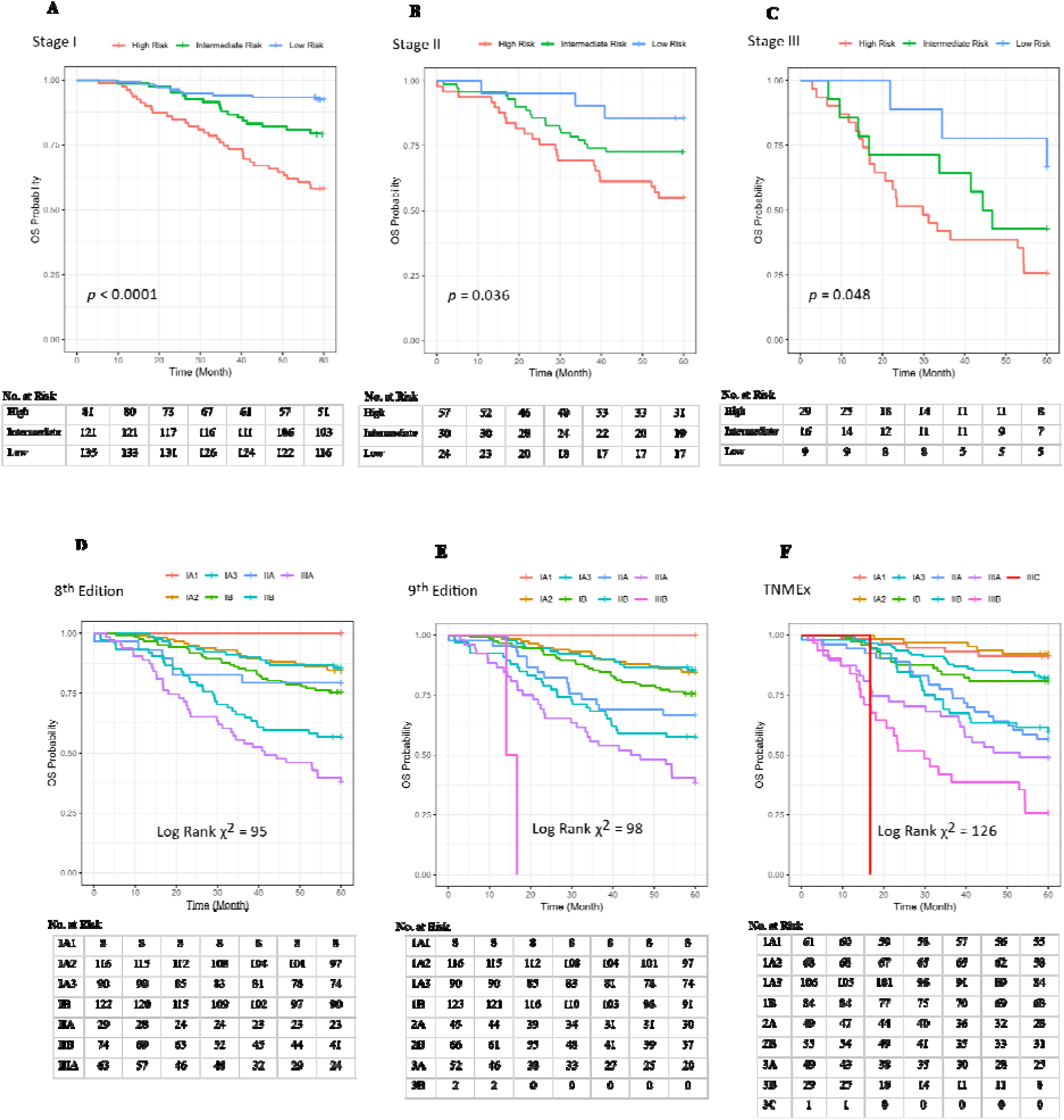
**(A-C)** Overall Survival Stratified by Molecular Risk Groups Within 9^th^ Edition TNM Stages in the Development Cohort (LORD, n = 502). **(D-F)** Overall survival of Lung Adenocarcinoma Patients Following Surgical Resection, Stratified by the 8^th^ Edition, 9^th^ Edition, and TNMEx staging systems in the Development Cohort (LORD, n = 502). Note: In the TNMEx model, Ex refers to incorporated gene expression profiles to the overall structure of the 9^th^ edition TNM staging system.

### 3.3. Validation and Prognostic Performance of TNMEx

TNMEx showed better separation of survival probabilities compared to conventional TNM stages **(Figure 2, D-F)**. The log-rank chi-square statistic was highest for TNMEx (χ² = 126) versus the 9^th^ (χ² = 96) and 8^th^ edition (χ² = 94). Multivariable Cox regression, showed the molecular risk score was found to be a predictor of OS independent of clinicopathological variables, with high-risk patients showing significantly poorer outcomes (HR = 2.53, 95% CI = 2.11 - 3.03, p = 0.0002) and intermediate-risk patients also demonstrating elevated risk (HR = 1.87, 95% CI = 1.39 - 2.31, p = 0.005), when compared to the low-risk group (**Tables 2,** and **S4**).

The TNMEx model showed stronger prognostic stratification for OS than the 9^th^ edition TNM, with higher hazard ratios comparing advanced versus early stages (TNMEx: HR = 4.56, 95% CI 4.12–4.91, p = 3.17×10 ¹² vs. 9^th^ edition TNM: HR = 3.24, 95% CI 2.71–4.04, p = 4.37×10□□). Furthermore, combining the molecular risk score with the 9^th^ edition TNM staging system (referred to as “9^th^ edition + risk score” in **Table 2**) yielded an even stronger prognostic model (HR = 4.87, 95% CI = 4.49 - 5.23, p = 1.11e-06), suggesting that risk stratification via gene expression provides substantial additive value beyond conventional staging **(Table 2**, and **S4)**. Calibration analysis using MAE and RMSE further supported the superior performance of TNMEx **(Table 2**, and **S5)**.

**Table 2:** Multivariable Cox Regression Analyses (with adjusting for other clinicopathological variables).

| Variable | HR (95%CI) | p-value | MAE | RMSE |
| --- | --- | --- | --- | --- |
| **Testing Group of the Development Cohort (LORD, n=152)** |  |  |  |  |
| **Risk category** (High risk) * | 2.53 (2.11–3.03) | **0.0002** | 0.11 | 0.15 |
| **Risk category** (Intermediate risk) * | 1.87 (1.39-2.31) | **0.005** | 0.13 | 0.16 |
| **8<sup>th</sup> edition** † | 2.95 (2.42-3.55) | **2.16e-05** | 0.17 | 0.21 |
| **9<sup>th</sup> edition** † | 3.24 (2.71-4.04) | **4.37e-09** | 0.12 | 0.15 |
| **TNMEx** † | 4.56 (4.12-4.91) | **3.17e-12** | 0.09 | 0.11 |
| **9<sup>th</sup> edition** †+ **risk score** | 4.87 (4.49-5.23) | **1.11e-06** | 0.10 | 0.13 |
| **Validation cohort (TCGA, n=271)** |  |  |  |  |
| **Risk category** (High risk) * | 2.49 (2.01-2.91) | **0.004** | 0.11 | 0.16 |
| **Risk category** (Intermediate risk) * | 1.51 (1.11-1.92) | **0.006** | 0.12 | 0.14 |

HR: hazard ratio, CI: confidence interval, MAE: Mean Absolute Error, RMSE: Root Mean Squared Error.
Note: In the TNMEx model, Ex refers to incorporated gene expression profiles to the overall structure of the 9<sup>th</sup> edition TNM staging system.
\* Compared with low-risk group.
† Stage II, III compared with stage I.
Bold text is used to highlight statistically significant p-values.

Model comparison metrics confirmed the superior performance of TNMEx in both testing and training cohort **(Table 3)**. Within the training cohort, TNMEx demonstrated a C-index of 0.73, outperforming the 9^th^ edition, which achieved 0.65 (p = 0.0001). Similarly, the AUC for TNMEx was 0.76, compared to 0.68 the 9^th^ edition (p = 0.002). Model fit, as evaluated by AIC, was also improved in TNMEx (AIC = 1113) versus the 9^th^ edition (AIC = 1127), indicating strong support for the improved model (ΔAIC > 10).

**Table 3:** Comparative performance of the 8^th^ and 9^th^ TNM editions and the TNMEx model in the training (n = 350) and testing (n = 152) groups of the LORD development cohort.

| Training Group of the Development Cohort (LORD, n=350) |  |  |  |  |  |  |  |  |  |
| --- | --- | --- | --- | --- | --- | --- | --- | --- | --- |
| Model | R<sup>2</sup> | C-Index | *p-value* | AUC | *p-value* | AIC | ΔAIC* | BIC | ΔBIC* |
| 8<sup>th</sup> Edition | 0.12 | 0.64 | 0.48<sup>a</sup><br>**0.0001<sup>b</sup>**<br>**0.0005<sup>c</sup>** | 0.67 | 0.39<sup>a</sup><br>**0.002<sup>b</sup>**<br>**0.0003<sup>c</sup>** | 1132 | 5<sup>a</sup><br>**14<sup>b</sup>**<br>**18<sup>c</sup>** | 1136 | 4<sup>a</sup><br>**15<sup>b</sup>**<br>**20<sup>c</sup>** |
| 9<sup>th</sup> Edition | 0.13 | 0.65 |  | 0.68 |  | 1127 |  | 1132 |  |
| TNMEx | 0.17 | 0.73 |  | 0.76 |  | 1113 |  | 1117 |  |
| 9<sup>th</sup> Edition + risk score | 0.19 | 0.74 |  | 0.79 |  | 1109 |  | 1112 |  |
| Testing Group of the Development Cohort (LORD, n=152) |  |  |  |  |  |  |  |  |  |
| Model | R<sup>2</sup> | C-Index | *p-value* | AUC | *p-value* | AIC | ΔAIC* | BIC | ΔBIC* |
| 8<sup>th</sup> Edition | 0.12 | 0.62 | 0.39<sup>a</sup><br>**0.006<sup>b</sup>**<br>**0.001<sup>c</sup>** | 0.67 | 0.27<sup>a</sup><br>**0.004<sup>b</sup>**<br>**0.001<sup>c</sup>** | 1130 | 1<sup>a</sup><br>**15<sup>b</sup>**<br>**19<sup>c</sup>** | 1135 | 1<sup>a</sup><br>**15<sup>b</sup>**<br>**21<sup>c</sup>** |
| 9<sup>th</sup> Edition | 0.12 | 0.65 |  | 0.67 |  | 1129 |  | 1134 |  |
| TNMEx | 0.17 | 0.72 |  | 0.75 |  | 1114 |  | 1119 |  |
| 9<sup>th</sup> Edition + risk score | 0.18 | 0.73 |  | 0.77 |  | 1110 |  | 1113 |  |

C-index: concordance index, AUC: area under the receiver operating characteristic curve, R<sup>2</sup> represents Royston's adaptation of Nagelkerke's R<sup>2</sup>, AIC: Akaike Information Criterion; BIC Bayesian Information Criterion.
Note: In the TNMEx model, Ex refers to incorporated gene expression profiles to the overall structure of the 9<sup>th</sup> edition TNM staging system.
a = The p-values refers to 8<sup>th</sup> Edition vs. 9<sup>th</sup> Edition TNM Stage in predicting survival.
b = The p-values refers to 9<sup>th</sup> Edition vs. TNMEx Stage in predicting survival.
c = The p-values refers to 9<sup>th</sup> Edition vs. 9<sup>th</sup> Edition plus risk scores in predicting survival.
\*A difference (Δ) in AIC or BIC of more than 2 suggests both models are equally good, a difference of 4-7 indicates moderate support for the model with the lower value, and a difference greater than 10 reflects strong preference for the model with the lower AIC or BIC.
Bold text is used to highlight statistically significant p-values.

When comparing the 8^th^ and 9^th^ editions of the TNM staging system, only marginal differences in prognostic performance were observed. The C-index increased slightly from 0.64 to 0.65 (p = 0.48), and the AUC improved minimally from 0.67 to 0.68 (p = 0.39), with no statistically significant difference. Similarly, reductions in AIC (1132 to 1127) and BIC (1136 to 1132) were modest, indicating that the 9^th^ edition did not provide substantial gains in prognostic accuracy over the 8^th^.

The TNMEx staging system from the training cohort was subsequently tested in the testing cohort. This reclassification system outperformed both the 8^th^ and 9^th^ editions, providing improved OS prediction. The C-index for TNMEx was 0.72, compared to 0.65 for the 9^th^ edition (p = 0.006). The AUC for TNMEx was 0.75, compared to 0.67 for the 9^th^ edition (p = 0.004). Model fit was also enhanced in TNMEx, with a lower AIC (1114) compared to the 9^th^ edition (AIC = 1129) and 8^th^ edition (AIC = 1130), supporting the robustness of the TNMEx system.

When comparing the 8^th^ and 9^th^ editions of the TNM staging system in the testing cohort, only minimal improvements were observed. The C-index increased modestly from 0.62 to 0.65, and the AUC remained essentially unchanged (0.67 for both editions), with no statistically significant differences (p = 0.39 and p = 0.27, respectively). Similarly, the reductions in AIC (1130 to 1129) was negligible, suggesting that the 9^th^ edition offered limited enhancement in prognostic discrimination relative to the 8^th^.

To further assess model performance, we evaluated reclassification and discrimination improvement metrics in the testing cohort. Compared with the 8^th^ edition, TNMEx demonstrated a significant net reclassification improvement (NRI = 0.27, 95% CI 0.18 - 0.35) and integrated discrimination improvement (IDI = 0.04, 95% CI = 0.01- 0.06). When compared with the 9^th^ edition, TNMEx showed even greater gains (NRI = 0.40, 95% CI 0.31- 0.47; IDI = 0.05, 95% CI = 0.02–0.08), whereas the 9^th^ edition itself did not meaningfully improve upon the 8^th^ (NRI = - 0.06, 95% CI - 0.09 to 0.01; IDI = 0.003, 95% CI = -0.02 - 0.02). These findings highlight the superior ability of TNMEx to reclassify patients into more appropriate prognostic groups, complementing its improved prognostic discrimination, calibration, and overall model fit **(Table 4)**.

**Table 4:**
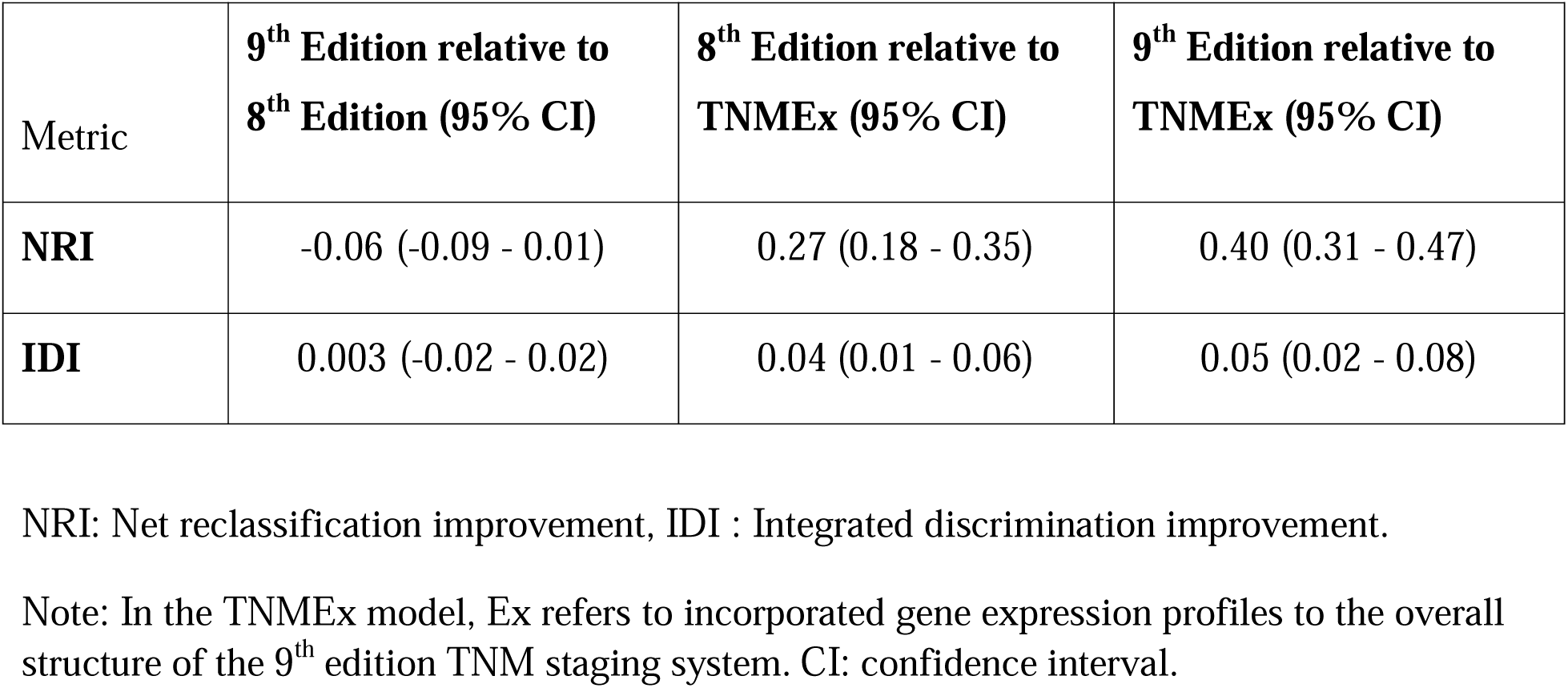
Comparative improvement metrics for the 8th edition, 9th edition, and TNMEx staging systems in the testing group of the development cohort (n = 152).

| Metric | **9<sup>th</sup> Edition relative to 8<sup>th</sup> Edition (95% CI)** | **8<sup>th</sup> Edition relative to TNMEx (95% CI)** | **9<sup>th</sup> Edition relative to TNMEx (95% CI)** |
| --- | --- | --- | --- |
| **NRI** | -0.06 (-0.09 - 0.01) | 0.27 (0.18 - 0.35) | 0.40 (0.31 - 0.47) |
| **IDI** | 0.003 (-0.02 - 0.02) | 0.04 (0.01 - 0.06) | 0.05 (0.02 - 0.08) |

NRI: Net reclassification improvement, IDI : Integrated discrimination improvement.
Note: In the TNMEx model, Ex refers to incorporated gene expression profiles to the overall structure of the 9<sup>th</sup> edition TNM staging system. CI: confidence interval.

### 3.4. External Validation Cohorts

We assessed the prognostic performance and generalizability of the developed molecular prognostic classifier in an independent cohort of 271 LUAD cases from TCGA. Of the 26 genes included in the prognostic classifier, 24 were available in the TCGA RNA-seq dataset **(Table S2,** and **S3)**. Using the same approach as in the development cohort, risk scores for each TCGA case were recalculated using the subset of 24 genes with available expression, applying the same LASSO-derived coefficients from the development cohort. Each patient received a molecular risk score, and stratification into low-, intermediate-, and high-risk groups was performed according to tertile thresholds. Molecular risk stratification distinguished OS outcomes, with high-risk patients showing significantly poorer survival than intermediate- and low-risk groups (HR high vs. low = 2.49, 95% CI 2.01–2.91, p = 0.004; HR intermediate vs. low = 1.51, 95% CI 1.11–1.92, p = 0.006). Predictive accuracy was also supported by calibration metrics, with MAE = 0.11 and RMSE = 0.16 for the high-risk group, and MAE = 0.12 and RMSE = 0.14 for the intermediate-risk group **(Table 2)**. Kaplan–Meier analysis validated the prognostic performance of the classifier in this independent dataset, with clear separation of survival curves across three molecular risk groups (p = 0.003) **(Figure S2)**. These findings demonstrate that the 26-gene signature retains prognostic value beyond the development LORD cohort, supporting its generalizability and robustness across independent populations, despite differences in staging editions used in TCGA.

To further assess the generalizability of the relative prognostic performance of the anatomical staging systems in an independent population, we evaluated the 8^th^ and 9^th^ edition TNM staging systems in an external cohort of 606 surgically resected LUAD patients from the NCCH. Kaplan–Meier analyses demonstrated significant survival differences across stages for both the 8th and 9th editions (log-rank p < 0.00001 for both), with comparable separation between stage groups **(Figure S3)**. The overall log-rank chi-square statistic indicated no meaningful improvement in stage discrimination between the 8^th^ and 9^th^ editions (χ² = 43 and χ² = 45, respectively). The C-index was identical for the 8^th^ and 9^th^ editions (both C-index = 0.73), with no significant difference in prognostic discrimination (p = 0.49). Likewise, AUC values were comparable between the 8th and 9th editions (0.67 vs. 0.68, p = 0.32), and model fit indices showed only marginal differences (AIC: 344 vs. 347; BIC: 353 vs. 358) **(Figure S5)**. These findings independently confirm that, despite statistically significant stage separation, the 9^th^ edition provides only limited incremental improvement over the 8^th^ edition in prognostic performance.

## 4. DISCUSSION

Despite continuous revisions to the TNM staging system, survival outcomes for patients with early-stage LUAD remain suboptimal [18–21, 38], with many high-risk patients not adequately identified by clinicopathologic parameters alone [1, 38–41]. Consistent with prior work on earlier editions [13, 27], our results show that integrating molecular classifiers significantly improves prognostic performance.

Using transcriptomic data from 502 resected LUAD cases, we developed a 26 gene molecular risk score that stratified patients into low-, intermediate-, and high-risk categories. These were incorporated into a novel staging model, TNMEx, which preserved the TNM hierarchy but allowed reclassification based on molecular risk. TNMEx outperformed both the 8^th^ and 9^th^ editions across multiple validation metrics, with calibration analyses showing superior alignment between predicted and observed survival.

Some of the genes incorporated in our prognostic algorithm are known elements of classical oncogenic pathways. Notably, six genes, *TMEM98* [42], *ABCC2* [43], *EPB41* [44], *H1-2* [45], *CASC9* [46], and *KYNU* [47], showed overlap with previously reported prognostic gene signatures in NSCLC, reinforcing the biological relevance of our model.

While the six overlapping genes have been previously linked to NSCLC prognosis, most of the remaining genes have not yet been characterized in lung cancer, despite their established or putative roles in oncogenic processes across other malignancies. This distinction highlights their potential as novel candidates for future investigation in LUAD.

Recent advances in gene-expression-based prognostic classifiers have demonstrated that transcriptomic programs capture biologic aggressiveness not reflected by anatomic staging alone. Notably, prior studies have developed molecular signatures such as the ORACLE classifier [23] and a validated 14-gene assay [48], which stratify risk within anatomically defined early-stage LUAD and identify patients with aggressive tumor biology despite favorable TNM stage. Building on this concept, two landmark studies further integrated a 14-gene expression signature into the 8^th^ edition TNM framework to create biologically informed staging systems for overall and recurrence-free survival, demonstrating improved prognostic discrimination while preserving the TNM structure [13, 27].

While these models established the feasibility and clinical value of molecularly informed staging, they were developed using earlier TNM editions. In contrast, our study represents the first integration of a transcriptomic prognostic classifier into the 9^th^ edition TNM staging system, reflecting contemporary staging definitions and current clinical practice. This extension is particularly relevant as the 9^th^ edition incorporates refined anatomic descriptors but still lacks a biological component, underscoring the need for complementary molecular stratification.

Importantly, the limited incremental prognostic value of the 9^th^ edition over the 8^th^ edition observed in the LORD cohort was independently confirmed in a large external cohort from Japan. Despite differences in ethnicity, healthcare systems, and clinical practice, the 9^th^ edition demonstrated only modest differences in prognostic performance compared with the 8^th^. This independent confirmation supports the generalizability of the current performance of anatomical staging systems and highlights the potential value of complementary biologically informed approaches such as TNMEx. Importantly, this observation does not diminish the clinical utility of anatomical staging, but rather underscores its limitations when used in isolation.

The clinical implications of our findings are notable. Accurate risk stratification in early-stage LUAD is critical, as approximately 13-23% of patients experience recurrence despite complete surgical resection [49]. Our classifier identifies these individuals and could influence postoperative treatment. As newer guidelines endorse risk-adapted therapy based on molecular and pathological features [50–52], models like TNMEx may offer a more personalized framework.

The TNMEx model provides a more nuanced risk stratification by up-staging high-risk and down-staging low-risk patients, potentially impacting adjuvant therapy decisions in borderline cases by directing treatment to those most likely to benefit while avoiding unnecessary therapy. For example, a high-risk stage IB patient under TNMEx may benefit from adjuvant chemotherapy despite being classified as low-risk under conventional staging, whereas a down-staged stage IIA patient may safely avoid additional adjuvant treatment.

Although combining the molecular risk score with the 9^th^ edition TNM staging system (“9^th^ edition + risk score”) yielded a strong prognostic model, the TNMEx framework offers greater simplicity and interpretability, providing a more practical and easily applicable tool for clinicians. By integrating a fourth component that reflects tumor biology, the TNMEx model remains practical for clinical use and provides a flexible structure that can be adapted to incorporate future molecular insights.

Findings were consistent in the external validation cohort obtained from the TCGA database. It is important to note that TNM staging information in the TCGA cohort was reported according to the 6^th^ or 7^th^ editions, rather than the most recent 9^th^ edition, and therefore we did not construct the TNMEx model in this dataset. Despite the absence of two genes in the TCGA dataset, the molecular prognostic classifier constructed using the remaining 24 genes consistently stratified patients by survival, supporting the robustness of the signature and suggesting that molecular classifiers can complement conventional staging across TNM editions and datasets.

One major strength of our study is the long follow-up period, which enabled robust assessment of long-term survival, a critical metric for evaluating staging performance. Another major strength is the use of bulk RNA sequencing, which captured the entire transcriptome rather than focusing on a limited set of genes. This comprehensive approach ensured that all potentially prognostic molecular signals were available for analysis, allowing for unbiased gene selection and the development of a robust classifier.

It is important to note that our analysis was performed on transcriptome-wide RNA sequencing data, which inherently captures the expression of all genes, including clinically actionable driver oncogenes such as *EGFR, KRAS, ALK, ROS1, BRAF, NTRK1/2/3, MET, RET,* and *ERBB2 (HER2)* [26, 53, 54]. Canonical drivers guide targeted therapies but are not consistently prognostic, mainly informing treatment rather than long-term survival. This study aimed to identify genes associated with overall prognosis, independent of therapy. The 26-gene LASSO signature reflects tumor aggressiveness, immune interactions, proliferation, and metastatic potential rather than druggable mutations. Nevertheless, combining actionable genomic alterations with transcriptomic signatures may further refine prognostic accuracy and enhance the clinical applicability of integrated molecular-TNM models.

However, the retrospective design may introduce selection bias, although the large sample size and long follow-up partially mitigate this limitation. In addition, the clinical implementation of RNA sequencing remains challenging, as it typically requires high-quality frozen tissue, which is not routinely available in diagnostic practice. Future efforts should therefore focus on developing targeted, clinically applicable gene panels suitable for formalin-fixed, paraffin-embedded (FFPE) specimens. While our findings support the integration of prognostic gene-expression profiles into staging systems, further validation in large, independent, multi-institutional cohorts is required. Finally, incorporating additional omics layers, such as mutational and epigenetic data, may further improve prognostic stratification.

An important consideration for the clinical implementation of invasive histologic descriptors such as histologic grade, STAS, LVI, and VPI is their interobserver reproducibility. Although formal interobserver agreement metrics were not assessed in this study, the use of standardized morphologic criteria and consensus review by specialized pulmonary pathologists supports the reproducibility of these histologic assessments.

## Conclusion

Across both the development and validation cohorts, incorporating our molecular prognostic classifier into the conventional TNM staging system consistently enhanced survival prediction, independent of the TNM edition used. In conclusion, our findings underscore the promise of a molecularly integrated staging system in LUAD. The TNMEx model represents a step toward biologically informed, personalized cancer staging, aligning with contemporary trends in precision oncology. Prospective, multicenter studies are warranted to validate and implement this approach in clinical practice.

## Funding sources

This work was supported by the IUCPQ Foundation, owing to a generous donation from Mr. Normand Lord. This work was also supported by the IUCPQ Biobank of the Quebec Respiratory Health research network.

## Data Availability

The data presented here are available from the corresponding author Philippe Joubert.

## Acknowledgments

The authors would like to thank all patients included in this study, and the IUCQP-UL Biobank, part of the Quebec Respiratory Health Research Network, for providing the clinical data.

## Notes

No Conflicts of Interest and no Source of Funding.

### Competing Interest Statement

The authors have declared no competing interest.

### Funding Statement

This study was funded by the Institut Universitaire de Cardiologie et de Pneumologie de Quebec-Laval University Foundation.

### Author Declarations

Ethics Approval and Consent to Participate: The study was approved by the ethics committee of the Institut Universitaire de Cardiologie et de Pneumologie de Quebec-Laval University, part of the Quebec Respiratory Health Research Network, approval number #2020-3352, 21871. The entire research was performed in accordance with the Declaration of Helsinki. Consent to Participate was waived due to retrospective type of study.

