## Supplementary Material for "Integration of a Molecular Prognostic Classifier into the Ninth Edition TNM Staging of Lung Adenocarcinoma"

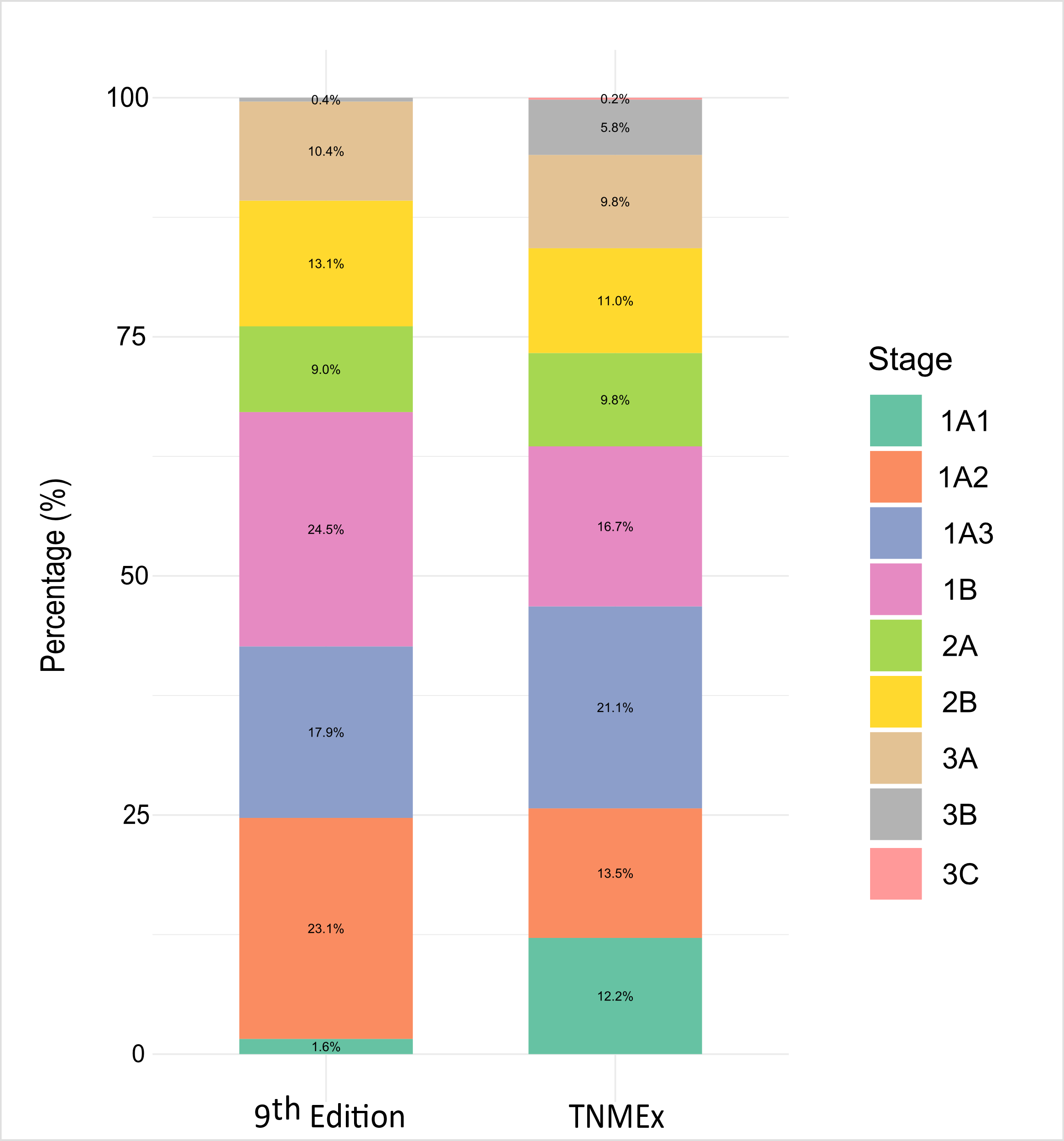

**Supplementary Figure 1:** Distribution of Patients Across Stages (1A1–3C) in the 9^th^ Edition TNM and TNMEx Staging Systems.

Note: In the TNMEx model, Ex refers to incorporated gene expression profiles to the overall structure of the 9^th^ edition TNM staging system.

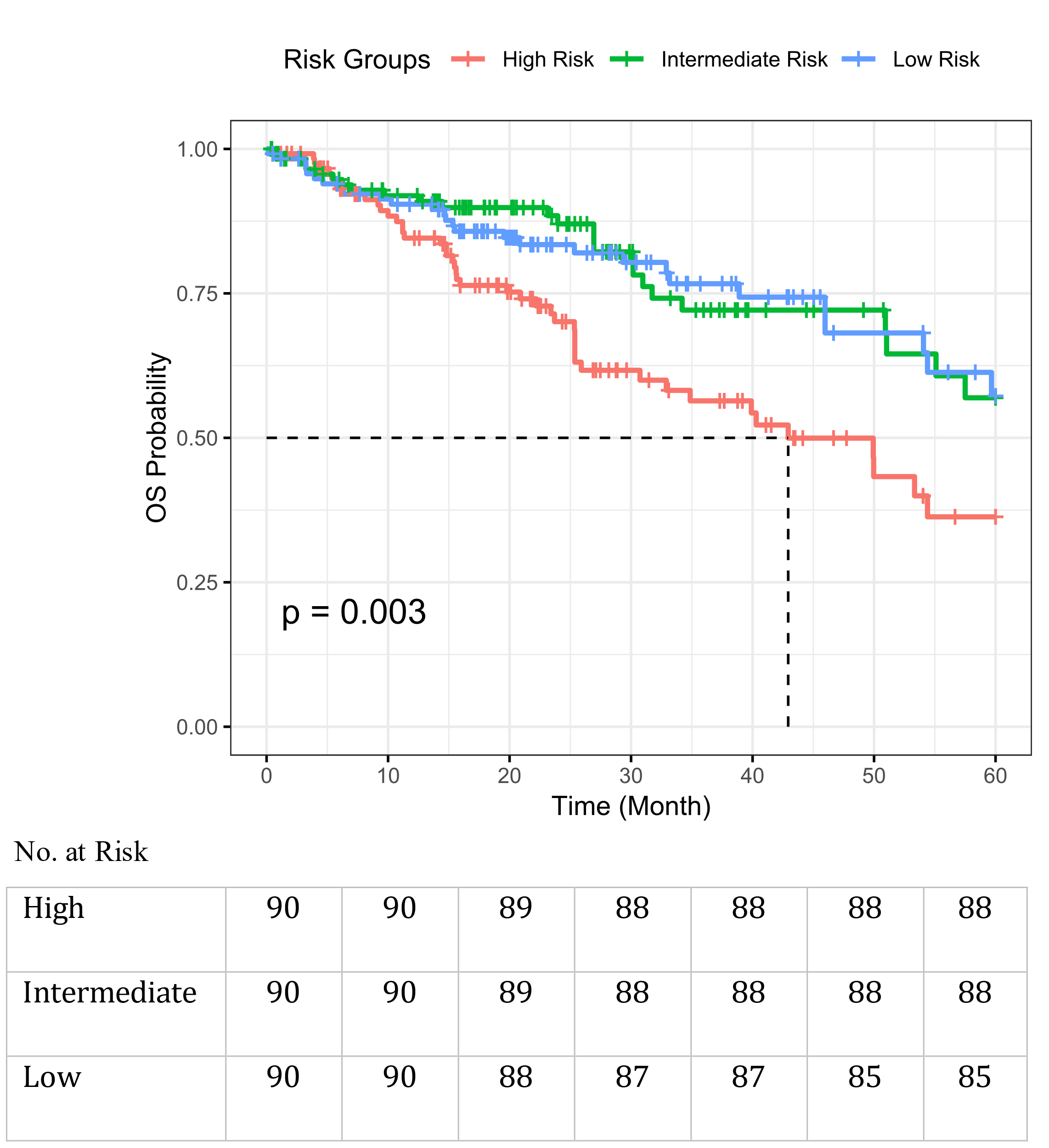

**Supplementary Figure 2:** Overall Survival Stratified by Molecular Risk Groups Within TCGA cohort. Kaplan-Meier curves demonstrate significant differences in overall survival (OS) probability between high-, intermediate-, and low-risk groups across. Risk scores were calculated per patient using a linear combination of gene expression levels weighted by log (HR) coefficients and used to stratify patients into low-, intermediate-, and high-risk groups based on tertiles of the score distribution.

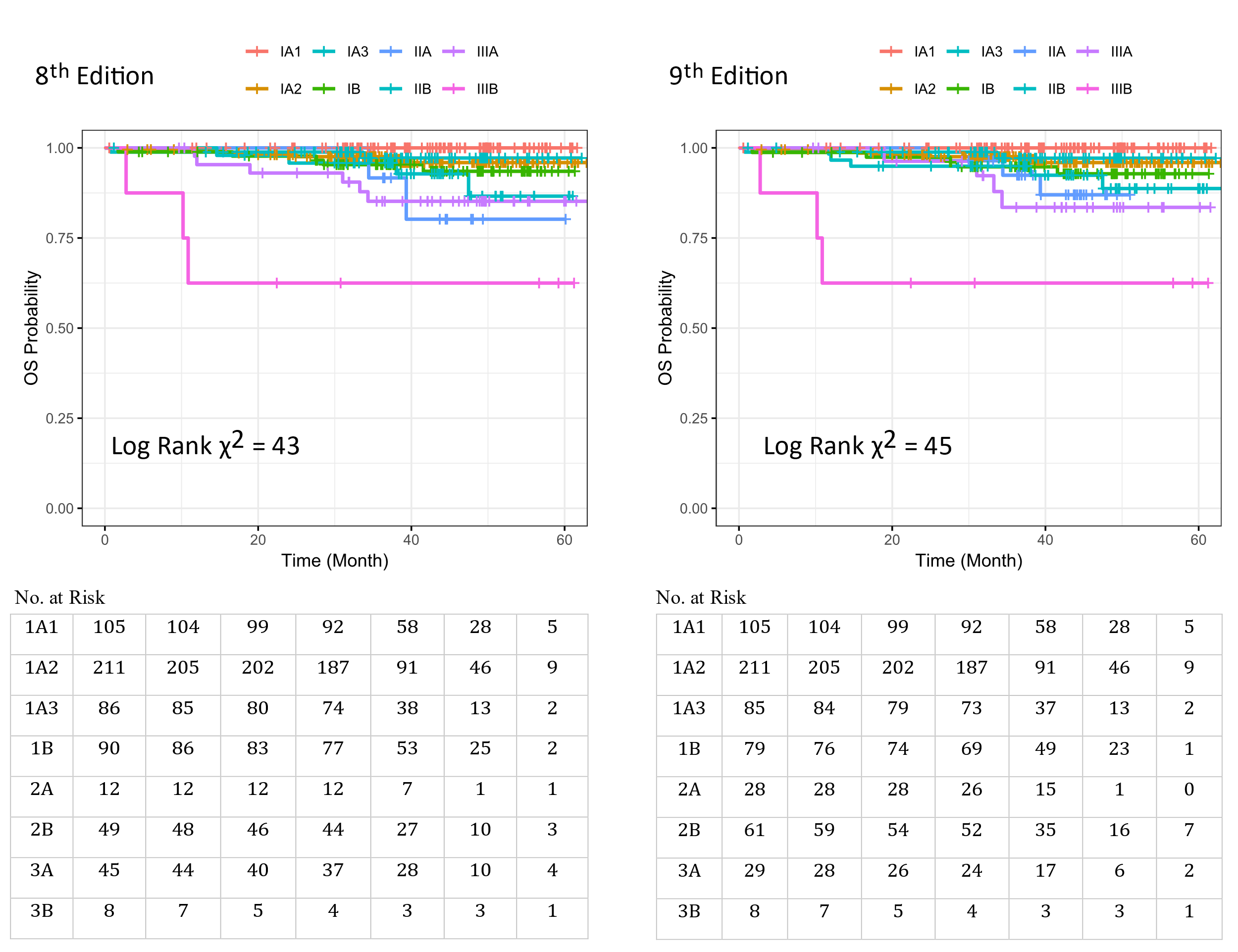

**Supplementary Figure 3:** Overall survival of Lung Adenocarcinoma Patients Following Surgical Resection, Stratified by the 8^th^ Edition,and 9^th^ Edition TNM staging systems in the Validation Cohort (NCCH, n = 606).

**Supplementary Table 1:** Overview of Patient and Tumor Characteristics in the Development Cohort (Training and Testing Groups)

| Characteristics | Training Group (n=350) | Testing Group (n=152) | p-value |
| --- | --- | --- | --- |
| Sex   - Female - Male | 190  160 | 85  67 | 0.735 |
| *Age at resection (year)*   - *Mean (SD)* - *Range* | *64.9 (8.1)*  *26-83* | *64.7 (8.5)*  *38-83* | 0.831 |
| Smoking Status   - Ever - Never | 327  23 | 145  7 | 0.393 |
| Pathologic N   - N0 - N1/2 | 275  75 | 120  32 | 0.924 |
| Pathologic T   - T1 - T2 - T3 - T4 | 173  138  23  16 | 76  58  13  5 | 0.789 |
| TNM Stage   - 1A1 - 1A2 - 1A3 - 1B - 2A - 2B - 3A - 3B | 7  81  60  86  34  42  38  2 | 1  35  30  37  11  24  14  0 | 0.679 |
| *Tumor size (cm)*   - *Mean (SD)* - *Median (Range)* | *2.9 (1.7)*  *2.4 (1.0-13.2)* | *2.9 (1.6)*  *2.5 (1.0-9.3)* | 0.769 |

**Supplementary Table 2:** List of Genes in the 26-Gene Molecular Prognostic Classifier

| **Ensemble** | **Gene ID** | **Protein location** | **Gene Symbol** | **Description** |
| --- | --- | --- | --- | --- |
| 1. ENSG00000006042 | 26022 | 17q11.2 | TMEM98 | Transmembrane protein 98 |
| 2. ENSG00000023839 | 1244 | 10q24.2 | ABCC2 | ATP binding cassette subfamily C member 2 |
| 3. ENSG00000101997 | 28952 | Xp11.23 | CCDC22 | Coiled-coil domain containing 22 |
| 4. ENSG00000111348 | 397 | 12p12.3 | ARHGDIB | Rho GDP dissociation inhibitor beta |
| 5. ENSG00000112561 | 7942 | 6p21.1 | TFEB | Transcription factor EB |
| 6. ENSG00000115325 | 1796 | 2p13.1 | DOK1 | Docking protein 1 |
| 7. ENSG00000115919 | 8942 | 2q22.2 | KYNU | kynureninase |
| 8. ENSG00000127124 | 59269 | 1p34.2 | HIVEP3 | HIVEP zinc finger 3 |
| 9. ENSG00000135148 | 10906 | 12q24.13 | TRAFD1 | TRAF-type zinc finger domain containing 1 |
| 10. ENSG00000142784 | 23038 | 1p36.11 | WDTC1 | WD and tetratricopeptide repeats 1 |
| 11. ENSG00000145632 | 10769 | 5q11.2 | PLK2 | Polo like kinase 2 |
| 12. ENSG00000156194* | 5470 | 4q21.1 | PPEF2 | Protein phosphatase with EF-hand domain 2 |
| 13. ENSG00000157303 | 203328 | 9q22.31 | SUSD3 | Sushi domain containing 3 |
| 14. ENSG00000159023 | 2035 | 1p35.3 | EPB41 | Erythrocyte membrane protein band 4.1 |
| 15. ENSG00000163866 | 113444 | 1p34.3 | SMIM12 | Small integral membrane protein 12 |
| 16. ENSG00000165175 | 58526 | Xp11.4 | MID1IP1 | MID1 interacting protein 1 |
| 17. ENSG00000181754 | 57463 | 1p13.3 | AMIGO1 | Adhesion molecule with Ig like domain 1 |
| 18. ENSG00000185100 | 122622 | 14q32.33 | ADSS1 | adenylosuccinate synthase 1 |
| 19. ENSG00000187837 | 3006 | 6p22.2 | H1-2 | H1.2 linker histone, cluster member |
| 20. ENSG00000198732 | 64093 | 14q24.2 | SMOC1 | SPARC related modular calcium binding 1 |
| 21. ENSG00000205744 | 79958 | 19p13.3 | DENND1C | DENN domain containing 1C |
| 22. ENSG00000213918 | 1773 | 16p13.3 | DNASE1 | Deoxyribonuclease 1 |
| 23. ENSG00000228288 | 641515 | 1q32.1 | MGAT4EP | MGAT4 family member E, pseudogene |
| 24. ENSG00000232093 | 100505666 | 1q21.3 | DCST1-AS1 | DCST1 antisense RNA 1 |
| 25. ENSG00000241631* | 106479337 | 17q11.2 | RN7SL316P | RNA, 7SL, cytoplasmic 316, pseudogene |
| 26. ENSG00000249395 | 101805492 | 8q21.13 | CASC9 | Cancer susceptibility 9 |

Gene annotations were obtained from public databases including the Ensembl Genome Browser (<https://www.ensembl.org>), GeneCards (<https://www.genecards.org>), and relevant literature.

*Not available in the TCGA cohort.

**Supplementary Table 3:** Functional Roles of Genes in the 26-Gene Prognostic Panel.

| **Gene** | **Ref** | **Gene Type** | **Function** |
| --- | --- | --- | --- |
| KYNU | [1] | Protein-coding | Mediates immune escape via the kynurenine pathway |
| DNASE1 | [2] | Protein-coding | Involved in apoptotic DNA fragmentation and chromatin degradation |
| ADSS1 | [3] | Protein-coding | Involved in purine biosynthesis; linked to metabolic stress tolerance |
| SMOC1 | [4] | Protein-coding | Inhibits TGF-β–driven angiogenesis; acts as tumor suppressor |
| AMIGO1 | [5] | Protein-coding | Regulates invasion via neural adhesion and microenvironment remodeling |
| TMEM98 | [6] | Protein-coding | Implicated in epithelial integrity and cell adhesion |
| ABCC2 | [7] | Protein-coding | Facilitates drug efflux; contributes to chemoresistance |
| CCDC22 | [8] | Protein-coding | Associated with NF-κB signaling and immune regulation |
| ARHGDIB | [9] | Protein-coding | Suppresses metastasis via Rho GTPase regulation |
| TFEB | [10] | Protein-coding | Controls autophagy and lysosomal function; aids survival under stress |
| DOK1 | [11] | Protein-coding | Tumor suppressor involved in MAPK signaling |
| HIVEP3 | [12] | Protein-coding | Regulates transcription of immune-related genes |
| TRAFD1 | [13] | Protein-coding | Inhibits toll-like receptor signaling; modulates tumor immune response |
| WDTC1 | [14] | Protein-coding | Involved in adipogenesis and possibly tumor metabolism |
| PLK2 | [15] | Protein-coding | Serine/threonine kinase involved in DNA damage response and cell cycle |
| PPEF2* | [16] | Protein-coding | Protein phosphatase; may regulate apoptosis |
| SUSD3 | [17] | Protein-coding | Emerging tumor suppressor in cell–cell signaling |
| EPB41 | [18] | Protein-coding | Suppresses Wnt/β-catenin signaling |
| SMIM12 | [19] | Protein-coding | Putative membrane signaling protein |
| MID1IP1 | [20] | Protein-coding | Regulates lipogenesis and metabolic adaptation |
| H1-2 | [21] | Protein-coding | Enhances antioxidant defense; promotes chemoresistance and tumor growth |
| DENND1C | [22] | Protein-coding | Links Rab35 activation to actin cytoskeleton |
| DCST1-AS1 | [23] | lncRNA | Promotes EMT and chemoresistance via TGF-β and AKT/mTOR signaling |
| CASC9 | [24] | lncRNA | Enhances proliferation and invasion through adhesion/migration pathways |
| MGAT4EP | [25] | Pseudogene | Pseudogene with pro-tumorigenic function |
| RN7SL316P* | [26] | Pseudogene | regulation of chromatin organization, nucleosome positioning, and nucleosomal DNA binding |

lncRNA: Long noncoding RNA

*Not available in the TCGA cohort.

**Supplementary Table 4:** Multivariable Cox-Regression Analysis for OS Entire Development Cohort (LORD, n=502).

| **Variable** | **HR (95% CI)** | **p-value** |
| --- | --- | --- |
| Risk category (High risk) * | 2.53 (2.11–3.03) | **0.0002** |
| Risk category (Intermediate risk) * | 1.87 (1.39-2.31) | **0.005** |
| 8^th^ Edition † | 2.95 (2.42-3.55) | **2.16e-05** |
| 9^th^ Edition † | 3.24 (2.71-4.04) | **4.37e-09** |
| TNMEx † | 4.56 (4.12-4.91) | **3.17e-12** |
| 9^th^ Edition †+ risk score | 4.87 (4.49-5.23) | **1.11e-06** |
| Age | 1.66 (1.42–2.31) | **0.030** |
| Sex female (vs. male) | 1.75 (1.58-1.95) | **0.021** |
| Smoking ever (vs. never) | 1.55 (0.93 - 1.92) | 0.065 |
| Grade 3 (vs. 1, 2) | 2.82 (2.40-3.18) | **0.001** |
| Tumor size | 1.87 (1.42-2.16) | **0.001** |
| LVI presence (vs. absence) | 2.27 (1.84-2.68) | **<0.0001** |
| VPI presence (vs. absence) | 1.22 (0.94-1.58) | 0.124 |
| STAS presence (vs. absence) | 1.63 (0.92-2.03) | 0.173 |

HR: hazard ratio, CI: confidence interval, LVI: lymphovascular invasion, STAS: spread through air spaces, VPI: visceral pleural invasion.

Note: In the TNMEx model, Ex refers to incorporated gene expression profiles to the overall structure of the 9^th^ edition TNM staging system.

* Compared with low-risk group.

† Stage II, III compared with stage I.

Bold emphasis is used to indicate statistically significant p-values.

**Supplementary Table 5:** Observed and Predicted Risks by Staging Models in the Development Cohort (LORD testing cohort, n=152 LUADs)

| **Stage** | **8^th^ Edition** | | | **9^th^ Edition** | | | **9^th^ Edition + Risk Score** | | | **TNMEx** | | |
| --- | --- | --- | --- | --- | --- | --- | --- | --- | --- | --- | --- | --- |
|  | **N** | **OR** | **PR** | **N** | **OR** | **PR** | **N** | **OR** | **PR** | **N** | **OR** | **PR** |
| **1A1** | 8 | 8.0 | 12.5 | 8 | 8.2 | 12.0 | 8 | 8.7 | 8.65 | 61 | 8.6 | 8.5 |
| **1A2** | 116 | 7.5 | 11.7 | 116 | 7.7 | 11.0 | 116 | 7.85 | 7.78 | 68 | 7.8 | 7.7 |
| **1A3** | 90 | 17.0 | 20.8 | 90 | 17.5 | 20.0 | 90 | 17.95 | 17.65 | 106 | 17.9 | 18.0 |
| **1B** | 122 | 18.2 | 23.0 | 123 | 18.8 | 22.0 | 123 | 19.3 | 18.9 | 84 | 19.2 | 19.0 |
| **2A** | 29 | 42.0 | 34.0 | 45 | 42.7 | 32.0 | 45 | 43.5 | 43.2 | 49 | 43.4 | 43.3 |
| **2B** | 74 | 40.1 | 36.5 | 66 | 40.7 | 35.0 | 66 | 40.5 | 40.1 | 55 | 40.4 | 40.3 |
| **3A** | 63 | 51.0 | 49.0 | 52 | 51.1 | 48.0 | 52 | 51.3 | 50.7 | 98 | 51.2 | 50.9 |
| **3B** | 0 | --- | --- | 2 | 46.9 | 42.0 | 2 | 47.1 | 46.0 | 29 | 47.0 | 46.0 |
| **3C** | 0 | --- | --- | 0 | --- | --- | 0 | --- | --- | 1 | 44.3 | 44.0 |

N: number of cases, OR: observed risk, PR: predicted risk.

| **Model** | **C-index** | **p-value** | **AUC** | **p-value** | **AIC** | **ΔAIC*** | **BIC** | **ΔBIC*** |
| --- | --- | --- | --- | --- | --- | --- | --- | --- |
| **8^th^ Edition** | 0.73 | 0.49 | 0.67 | 0.32 | 344 | 3 | 353 | 5 |
| **9^th^ Edition** | 0.73 |  | 0.68 |  | 347 |  | 358 |  |

**Supplementary Table 5:** Comparative performance of the 8^th^ and 9^th^ TNM editions in the NCCH cohort (n = 607).

C-index: concordance index, AUC: area under the receiver operating characteristic curve, AIC: Akaike Information Criterion; BIC Bayesian Information Criterion.

*A difference (Δ) in AIC or BIC of more than 2 suggests both models are equally good, a difference of 4-7 indicates moderate support for the model with the lower value, and a difference greater than 10 reflects strong preference for the model with the lower AIC or BIC.

**Supplementary Methods 1:** Gene Expression Quantification

Total RNA was extracted using the RNeasy Universal Plus Mini Kit with Qiazol lysis reagent (Qiagen, Hilden, Germany) according to the manufacturer’s protocol. mRNA libraries were prepared using the Illumina Stranded mRNA Prep Ligation Kit (Illumina, San Diego, CA, USA). High-throughput sequencing was performed at Genome Québec (Montreal, QC, Canada) using the Illumina NovaSeq 6000 platform, aiming for a minimum coverage of 75 million paired-end reads (2 × 100 bp) per sample. In total, RNA from tumor tissue of 502 patients was sequenced across five independent batches. Transcript quantification was performed using a pseudo-alignment method with KALLISTO (Bray et al., 2016), which assigns k-mer reads directly to transcript sequences based on the GENCODE v44 reference transcriptome.

The initial expression matrix included both protein-coding and non-coding genes (lncRNA, pseudogene, and antisense). Transcript-level data were then aggregated to the gene level using the tximport and AnnotationDbi R packages, resulting in expression estimates for 62,700 genes, reported in both estimated counts and TPM (transcripts per million) values.

**Supplementary Methods 2:** Statistical Analyses

The normalized expression values of the top 20% most variable genes (12,540 out of 62,700) were used to retain genes contributing the most to transcriptomic variation across samples. To assess the stability of our approach, we evaluated multiple gene-selection thresholds, beginning with the top 1% most variable genes and increasing in 5% intervals, as well as subsets defined by high expression levels. Because these different strategies yielded nearly identical performance, we adopted the top 20% most variable genes as a representative and reliable selection for subsequent analyses [27, 28]. Prior to model construction, gene expression values were variance-stabilizing transformed (vst) and z-score normalized to ensure comparability across samples. Although variance filtering alone may include some biologically less informative genes, LASSO-penalized regression analyses further refined the feature set to prognostically relevant genes, thereby enhancing biological specificity and reducing noise propagation in model construction [29].

The LASSO-penalized Cox regression was implemented using the *glmnet* package (V4.1.8). The cv.glmnet() function was applied with parameters family = "cox", alpha = 1, and maxit = 100000 using OS as the primary endpoint. The optimal penalty parameter (λ) was determined through repeated 10-fold cross-validation to prevent overfitting, ensuring that model performance was evaluated on unseen data during each iteration. This cross-validation procedure was repeated 100 times with different random seeds to enhance model robustness and assess feature stability. Gene selection frequencies were computed across all iterations, and features consistently retained in a high proportion of runs (>70%) were considered stable, demonstrating the robustness and reproducibility of the molecular classifier. Genes with non-zero coefficients at lambda.min that were consistently retained across iterations were extracted using the coef() function and carried forward for downstream analysis. The 1-standard error rule (1-SE) was applied to favor a more parsimonious model.

The functional information of the genes was obtained using the *Metascape, KEGG, and Gene Ontology (GO)* databases, which provide gene annotation and enrichment analysis. Additional insights were gathered through a manual review of relevant scientific literature. The prognostically relevant genes were used to generate the molecular prognostic classifier. To construct a molecular risk score for each patient, we calculated each patient’s score by combining the selected gene expression values, multiplying each one by its coefficient obtained from the LASSO-Cox model. The model assumes a linear relationship between normalized gene expression and log hazard; this assumption was assessed using Schoenfeld residuals and by inspecting partial residual plots, confirming adequate linearity.

Survival analyses were conducted using Kaplan-Meier curves and multivariable Cox regression models, implemented with the *survival* (V3.6.4) and *survminer (V0.5.0)* R packages. A multivariable Cox regression analysis was performed to assess the independent prognostic significance of the molecular classifier, controlling for clinicopathological factors including age, sex, smoking status, tumor grade, TNM stage, tumor size, STAS, LVI, and VPI.

To assess the added prognostic value of the molecular classifier, we compared the performance of three staging models: the 8^th^ edition TNM staging, the 9^th^ edition TNM staging, and the integrated TNMEx model. The performance of the model was assessed using hazard ratios (HRs), concordance index (C-index), area under the curve (AUC), chi-square statistics, Akaike Information Criterion (AIC), and Bayesian Information Criterion (BIC), along with Nagelkerke’s R² for each staging to assess model fit. The mean absolute error (MAE) and root mean squared error (RMSE) were computed to quantify prediction accuracy between observed and predicted survival probabilities. We also assessed the added predictive benefit of the TNMEx model over the conventional 8^th^ and 9^th^ edition TNM staging systems, by computing the Net Reclassification Improvement (NRI) and the Integrated Discrimination Improvement (IDI). NRI measures the improvement in correctly reclassifying patients into higher- or lower-risk categories, while IDI reflects the average increase in model discrimination by comparing the differences in mean predicted probabilities between models.

To ensure robust model development and generalizability, the LORD cohort (n = 502) was randomly partitioned into a training cohort (n = 350) and an independent testing cohort (n = 152) using a 3:1 ratio, ensuring balanced distribution of key clinicopathological variables between the two groups **(Table S1)**. The training set served for model construction and internal optimization, while the testing set was reserved for independent internal validation. An additional external validation cohort consisting of 271 LUAD cases from TCGA was used to confirm the model’s reproducibility and generalizability across populations. This three-tier validation framework allowed comprehensive evaluation of performance across independent datasets. All statistical analyses were performed in R (version 4.5.1, RStudio, Boston, MA, USA). P-values were two-sided, with significance defined as p < 0.05 unless specified otherwise.

**Supplementary Methods 3:** Molecular Prognostic Classifier, and Novel TNM Staging Development

We developed a molecular prognostic classifier and integrated into the 9^th^ edition TNM staging system to improve risk stratification and survival prediction in LUAD. To identify the most prognostically informative genes in tumor samples, we applied a Least Absolute Shrinkage and Selection Operator (LASSO) penalized Cox regression model, using 5-years OS as primary endpoint. Then a molecular risk score for each patient was calculated.

Patients were subsequently categorized into low-, intermediate-, and high-risk groups based on tertiles of the molecular risk score: the lowest 33^rd^ percentile defined the low-risk group, the highest 33^rd^ percentile defined the high-risk group, and those in between were assigned to the intermediate-risk group. This approach ensured balanced sample sizes across risk categories allowing for the delineation of prognostic subgroups based on gene expression profiles.

Then the molecular risk grouping was incorporated into the 9^th^ edition TNM staging system to develop a novel integrated staging model, referred to as TNMEx, where “Ex” represents gene expression profiles. The reclassification process was guided by molecular risk group assignments: high-risk patients were up-staged by one TNM level, low-risk patients were down-staged by one level, and intermediate-risk patients remained in their original stage. For example, a stage IA3 tumor in a high-risk patient would be reclassified as stage IB, whereas a stage IA3 tumor in a low-risk patient would be down-staged to stage IA2.

We also explored a more granular molecular stratification using five risk groups (very low, low, intermediate, high, and very high). In this extended model, very high-risk tumors were up-staged by two TNM levels and very low–risk tumors were down-staged by two levels, while low- and high-risk groups were shifted by one level as in the primary system. However, this five-tier approach did not enhance prognostic performance relative to the original three-tier model (data not shown). For this reason, and to maintain clinical simplicity and interpretability, we adopted the three-group molecular classifier and its corresponding one-level adjustment scheme for TNMEx.

**Supplementary Methods 4:** Definitions of Performance Metrics and LASSO Model Details

For a given patient i, the risk score is computed as:

***Risk Scoreᵢ = Σ (βⱼ × xᵢⱼ)***

Where xᵢⱼ is the log2-transformed, standardized expression of gene j in patient I, βⱼ is the coefficient (log hazard ratio) for gene j, estimated by the LASSO-penalized Cox regression, and p is the number of genes selected by the model (non-zero coefficients at λ_min).

The LASSO penalty is applied during model fitting, and the objective function minimized is:

***L(β) = – ℓ(β) + λ × Σ |βⱼ|***

Where ℓ(β) is the partial log-likelihood of the Cox model, λ is the penalty parameter determined by cross-validation, and |βⱼ| represents the L1 norm, which encourages sparsity by shrinking some coefficients to zero.

Mean Absolute Error (MAE) per staging system:

***MAE = (1/n) × Σ |predicted risk − observed risk|***

Root Mean Squared Error (RMSE):

***RMSE = √[(1/n) × Σ (predicted risk − observed risk)***
